## Supplementary Information for "Risk and rates of hospitalisation in young children: a prospective study of a South African birth cohort"

**This PDF file includes:**

**Supplementary Figure 1:** DAG model

**Supplementary Figure 2**. Causes of hospitalisation stratified by HIV exposure and age groups

**Supplementary Table 1:** Incidence of hospitalisations in the first 2 years of life stratified by age

**Supplementary Table 2:** Proportion of LRTI and RSV-LRTI hospitalisations by age category

**Supplementary Table 3:** Incidence of hospitalisations in the first 2 years of life by HIV exposure status excluding recurrent events

**Supplementary Table 4:** Incidence of hospitalisations in the first 2 years of life by HIV exposure status

**Supplementary Table 5:** Impact of birth, breastfeeding, and HIV-related factors on hospitalisation, stratified by HIV status and age

**Supplementary Table 6:** Association between malnutrition and hospitalisation

**Supplementary Figure 1:** DAG model
Directed Acyclic Graph (DAG) constructed in www.dagitty.net: HIV exposure and child hospitalisation. This DAG was constructed to examine for possible confounding in the relationship between HIV exposure and child hospitalisation from 0 -24 months in the Drakenstein Child Health Study, South Africa.

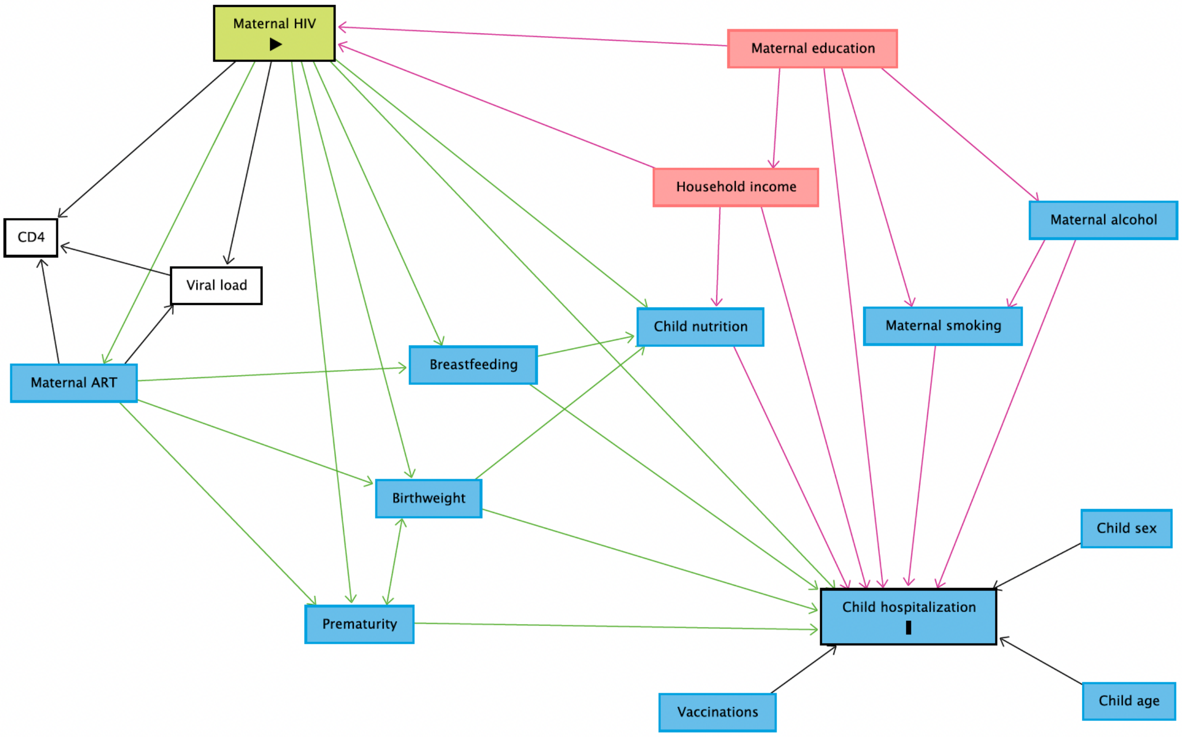


***Footnote:*** Minimal sufficient adjustment sets for estimating the total effect of maternal HIV exposure on child hospitalisation include: Household income and maternal education.

**Legend**

- exposure
- outcome
- ancestor of exposure
- ancestor of outcome
- ancestor of exposure *and* outcome
- adjusted variable
- unobserved (latent)
- other variable
- causal path
- biasing path

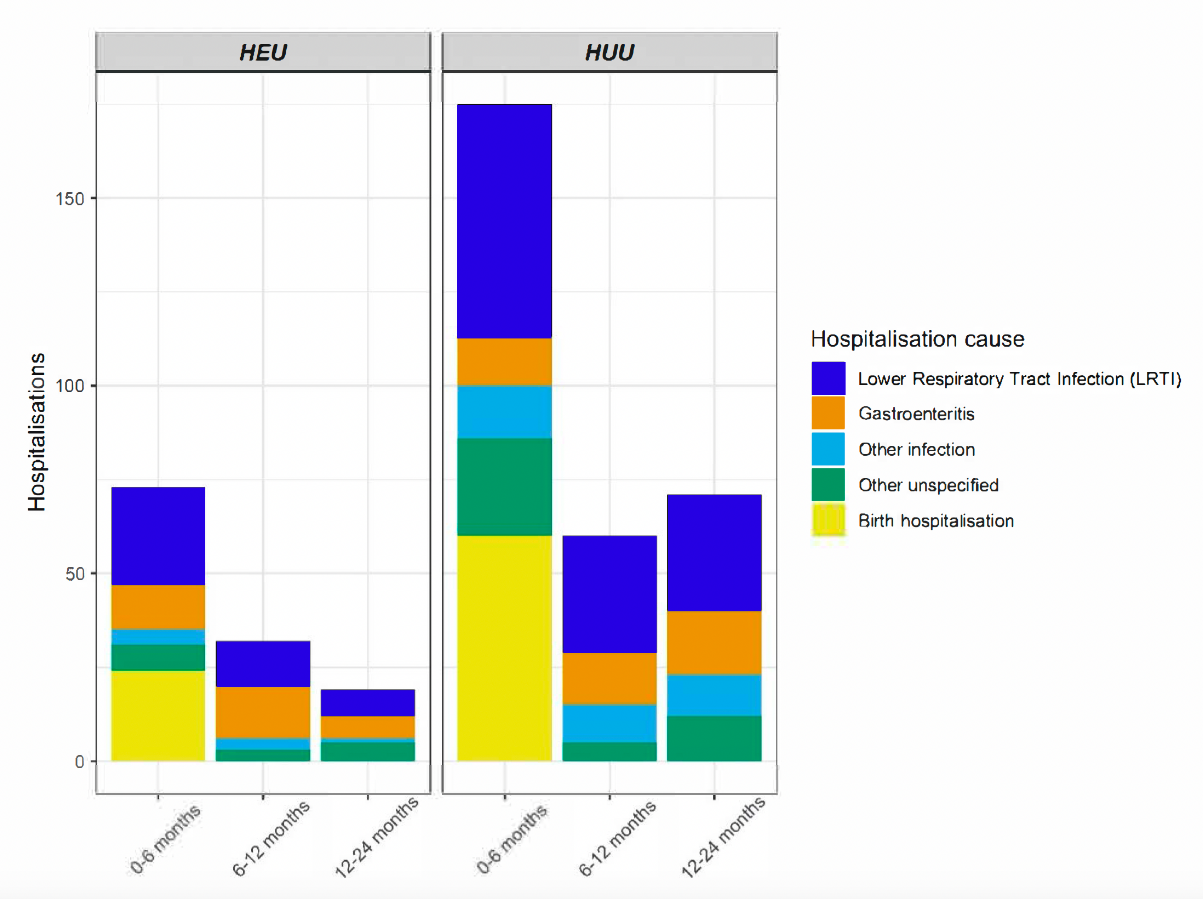


**Supplementary Figure 2**. Causes of hospitalisation stratified by HIV exposure and age groups
Other infections include: meningitis, sepsis, otitis media, urinary tract infection. Other unspecified causes include: Accident and trauma, burns, seizures, nutritional issues (failure to thrive, protein energy malnutrition, micronutrient deficiencies including anaemia)

**Supplementary Table 1**: Incidence of hospitalisation in the first 2 years of life stratified by age

| | IR / 1000 person years (95% CI) | IRR (95% CI) |
|---|---|---|
| *All hospitalisations* | | |
| 0-12 months | 314 (281-349) | *0-12 v 12-24 months:* 3.81 (2.99-4.85) * |
| 0-6 months | 454 (399-514) | *0-6 v 6-12 months*: 2.69 (2.11-3.44) * |
| 6-12 months | 168 (135-207) | *6-12 v 12-24 months:* 2.04 (1.51-2.76) * |
| 12-24 months | 82 (65-102) | *0-6 v 12-24 months*: 5.51 (4.30-7.70) * |

*Footnote*: IR = Incidence rate; IRR = Incidence rate ratio; HR = Hazard ratio. *p-value < 0.001

**Supplementary Table 2:** Proportion of LRTI and RSV-LRTI hospitalisations by age category

| | LRTI hospitalizations (n; % of total LRTI hospitalisations [n=169]) | RS -LRTI (n; % of total RSV hospitalisations [n=53]) | LRTI hospitalisations (n; % of hospitalisations per age group) | RSV-LRTI (n; % of hospitalisations per age group) |
|---|---|---|---|---|
| **Age category** | | | | |
| **0-12 months** | 131/169 (78%) | 46/53 (87%) | 131/256 (51%) | 46/256 (18%) |
| **0-6 months** | 88/169 (52%) | 36/53 (68%) | 88/164 (54%) | 36/164 (22%) |
| **6-12 months** | 43/169 (25%) | 10/53 (19%) | 43/92 (47%) | 10/92 (11%) |
| **12-24 months** | 38/169 (22%) | 7/53 (13%) | 38/90 (42%) | 7/90 (8%) |

*Footnote:* Numbers exclude birth hospitalisations. Abbreviations: LRTI: Lower respiratory tract infection; RSV: Respiratory syncytial virus

**Supplementary Table 3**: Incidence of hospitalisations in the first 2 years of life by HIV exposure status excluding recurrent events

| | **HEU IR / 1000 person years** | **HUU IR /1000 person years** | **IRR (95% CI)** |
|---|---|---|---|
| <u>*All hospitalisations*</u> | | | |
| 0-12 months | 334 (262-418) | 223 (192-258) | 1.49 (1.14-1.95) ** |
| 0-6 months | 510 (388-658) | 349 (295-410) | 1.46 (1.08-1.97) * |
| 6-12 months | 144 (82-234) | 89 (62-123) | 1.62 (0.90-2.92) |
| 12-24 months | 50 (25-90) | 45 (31-62) | 1.12 (0.57-2.20) |

*Footnote:* ** p-value < 0.01; * p-value < 0.05. Abbreviations: HEU = HIV-exposed uninfected; HUU = HIV-unexposed uninfected; IR = Incidence rate; IRR = Incidence rate ratio

**Supplementary Table 4:** Incidence of hospitalisations in the first 2 years of life by HIV exposure status

| | | | | *Unadjusted* | *Adjusted model 1* | *Adjusted model 2* |
|---|---|---|---|---|---|---|
| | **All IR / 1000 person years** | **HEU IR / 1000 person years** | **HUU IR /1000 person years** | **IRR (95% CI)** | **HR (95% CI)** | **HR (95% CI)** |
| 0-12 months | 235 (207-266) | 346 (274-431) | 205 (175-238) | 1.69 (1.30-2.21) *** | 1.53 (1.14-2.07) ** | 1.59 (1.16-2.18) ** |
| 0-6 months | 299 (255-349) | 424 (314-559) | 265 (219-319) | 1.60 (1.14-2.23) ** | 1.48 (1.03-2.12) * | 1.56 (1.07-2.28) * |
| 6-12 months | 168 (135-207) | 265 (179-379) | 142 (107-183) | 1.87 (1.20-2.91) ** | 1.73 (1.05-2.85) * | 1.76 (1.00-3.08) * |
| 12-24 months | 82 (65-102) | 77 (45-124) | 84 (65-106) | 0.92 (0.54-1.57) | 0.91 (0.53-1.55) | 0.78 (0.44-1.38) |

*Footnote*: Unadjusted incident rate ratios and adjusted hazard models for hospitalisations in the first 2 years of life by HIV exposure excluding birth hospitalisations.
IR = Incidence rate; IRR = Incidence rate ratio; HR = Hazard ratio. Multivariate models adjusted for (1) maternal education and household income; (2) maternal education, household income, maternal age at birth and maternal smoking. *** p-value < 0.001; ** p-value < 0.01; * p-value < 0.05
Abbreviations: HR = Hazard ratio; HEU = HIV-exposed uninfected; HUU = HIV-unexposed uninfected; IR = Incidence rate; IRR = Incidence rate ratio; OR = Odds ratio.

**Supplementary Table 5:** Impact of birth, breastfeeding and HIV-related factors on hospitalisations stratified by HIV status and age

| | | HEU<br>HR (95% CI) | HUU<br>HR (95% CI) | Total<br>HR (95% CI) |
|---|---|---|---|---|
| ***Birth factors*** | | | | |
| *Prematurity* | | | | |
| 0-12 months | | 1.94 (1.11-3.37) * | 1.75 (1.22-2.52) ** | 1.85 (1.36-2.52) *** |
| | 0-6 months | 1.67 (0.89-3.13) | 2.04 (1.34-3.11) *** | 1.95 (1.37-2.78) *** |
| | 6-12 months | 2.66 (1.07-6.62) * | 1.26 (0.66-2.41) | 1.74 (0.99-3.04) |
| 12-24 months | | 1.48 (0.43-5.11) | 0.97 (0.45-2.06) | 1.07 (0.56-2.04) |
| ***Feeding*** | | | | |
| *Ever breastfed* | | | | |
| 0-12 months | | 0.85 (0.51-1.43) | 0.76 (0.42-1.39) | 0.63 (0.46-0.87) ** |
| 12-24 months | | 1.00 (0.39-2.62) | 1.07 (0.35-3.22) | 1.08 (0.60-1.94) |
| *Breastfeeding for a year* | | | | |
| 0-12 months | | 1.02 (0.39-2.62) | 0.71 (0.50-1.02) | 0.67 (0.48 -0.93) * |
| 12-24 months | | 0.97 (0.24-3.88) | 0.64 (0.36-1.11) | 0.70 (0.42 -1.16) |
| *Immunization timing by 9 months* | | | | |
| 0-12 months | | 1.48 (0.87-2.52) | 1.36 (0.96-1.92) | 1.39 (1.04-1.87) * |
| 12-24 months | | 1.34 (0.52-3.48) | 1.03 (0.60-1.77) | 1.09 (0.68-1.75) |
| ***HIV-related variables*** | | | | |
| *CD4 - categorical* | | | | |
| *(>500 vs <=500)* | | | | |
| 0-12 months | | 0.99 (0.56-1.76) | - | - |
| | 0-6 months | 0.92 (0.46-1.87) | - | - |
| | 6-12 months | 1.11 (0.43-2.83) | - | - |
| 12-24 months | | 0.88 (0.33-2.35) | - | - |
| *Viral load-categorical* | | | | |
| *(>= 40 vs <40)* | | | | |
| 0-12 months | | 1.51 (0.79-2.86) | - | - |
| | 0-6 months | 0.92 (0.43-1.98) | - | - |
| | 6-12 months | 4.43 (1.52-12.87) ** | - | - |
| 12-24 months | | 1.31 (0.40-4.30) | - | - |
| *ART regimen initiation* | | | | |
| *(Before vs during pregnancy)* | | | | |
| 0-12 months | | 0.86 (0.51-1.44) | - | - |
| | 0-6 months | 1.06 (0.58-1.93) | - | - |
| | 6-12 months | 0.54 (0.20-1.40) | - | - |
| 12-24 months | | 0.76 (0.29-1.99) | - | - |

*Footnote:* Results with birth hospitalisations excluded. *** p-value < 0.001; ** p-value < 0.01; * p-value < 0.05 for unadjusted models. Definitions: Prematurity (<37 vs >= 37 weeks); Ever breastfeeding (>0 months vs = 0 months); Breastfeeding for a year (>11 months vs <= 11 months). Abbreviations: ART = antiretroviral therapy; HR = Hazard ratio; HEU = HIV-exposed uninfected; HUU = HIV-unexposed uninfected

**Supplementary Table 6:** Association between malnutrition and hospitalisation

| | Underweight | | | |
|---|---|---|---|---|
| | **Total** | **HEU** | **HUU** | **OR (95% CI)** |
| 0-12 months | 47 (19%) | 19 (24%) | 28 (17%) | 1.62 (0.83-3.12) |
| 0-6 months | 35 (22%) | 14 (29%) | 21 (19%) | 1.76 (0.80-3.85) |
| 6-12 months | 12 (14%) | 5 (17%) | 7 (12%) | 1.46 (0.40-5.03) |
| 12-24 months | 9 (11%) | 4 (25%) | 5 (7.6%) | 4.07 (0.90-17.69) |
| | **Stunted** | | | |
| 0-12 months | 71 (31%) | 27 (38%) | 44 (28%) | 1.63 (0.90-2.94) |
| 0-6 months | 47 (32%) | 16 (38%) | 31 (30%) | 1.45 (0.68-3.06) |
| 6-12 months | 24 (28%) | 11 (38%) | 13 (23%) | 2.07 (0.78-5.51) |
| 12-24 months | 14 (18%) | 4 (27%) | 10 (16%) | 2.41 (0.65-8.32) |
| | **Wasted** | | | |
| 0-12 months | 29 (15%) | 11 (19%) | 18 (14%) | 1.50 (0.64-3.37) |
| 0-6 months | 18 (17%) | 8 (28%) | 10 (13%) | 2.51 (0.86-7.22) |
| 6-12 months | 11 (13%) | 3 (10%) | 8 (14%) | 0.71 (0.15-2.68) |
| 12-24 months | 8 (10%) | 3 (20%) | 5 (7.9%) | 2.90 (0.54-13.55) |

Abbreviations: HEU = HIV-exposed uninfected; HUU = HIV-unexposed uninfected; OR = Odds ratio
